# Left hemisphere lateralization of epileptic focus can be more frequent in temporal lobe epilepsy surgical patients with no consensus associated with depression lateralization

**DOI:** 10.1101/2020.08.20.20178244

**Authors:** Graciane Radaelli, Fernanda Majolo, Eduardo Leal-Conceição, Francisco de Souza Santos, Vinícius Escobar, Gabriele Goulart Zanirati, Mirna Wetters Portuguez, Fulvio Alexandre Scorza, Jaderson Costa da Costa

## Abstract

Temporal Lobe Epilepsy (TLE) is considered to be the most common form of epilepsy, and it has been seen that most patients are refractory to antiepileptic drugs. A strong association of this ailment has been established with psychiatric comorbidities, primarily mood and anxiety disorders. The side of epileptogenic may contribute to depressive and anxiety symptoms; thus, in this study, we performed a systematic review to evaluate the prevalence of depression in TLE in surgical patients. The literature search was performed using PubMed/Medline, Web of Science, and PsycNET to gather data from inception until January 2019. The search strategy was related to temporal lobe epilepsy, depressive disorder and anxiety. After reading full texts, 14 articles meeting the inclusion criteria were screened. The main method utilized for psychiatric diagnosis was DSM/SCID. However, most studies failed to perform the neuropsychological evaluation. For those with lateralization of epileptic focus mostly occurred in the left hemisphere. For individual depressive diagnosis, nine studies were evaluated and five for anxiety. Therefore, from the data analyzed in both situations, no diagnosis was representative in preoperative and postoperative cases. In order to estimate the efficacy of surgery in the psychiatry episodes and its relation to seizure control, the risk of depression and anxiety symptoms in epileptic patients need to be determined before surgical procedures. Rigorous pre- and postoperative evaluation is essential for psychiatry conditions in patients with refractory epilepsy candidates for surgery.

## Introduction

The most common form of focal epilepsy is Temporal Lobe Epilepsy (TLE). Most of the patients are refractory to antiepileptic drugs (AEDs), about 75% presenting resistance to medical treatment [1–3]. Psychiatric comorbidities, such as anxiety disorders, are often expressed among patients with TLE [4–7]. The exact mechanism behind this association remains to be explored; however, the major findings indicate that these comorbidities and TLE share similar neuroanatomic localization [3, 8]. Another factor that remains uncertain and may contribute to anxiety and depressive symptoms in TLE is the side of the epileptogenic focus. Research has documented the prevalence of interictal depression in left-sided seizure foci [9, 10], while others illustrate a tendency for greater depressive symptoms in presurgical right-sided seizure foci patients [11, 12].

Depressive and anxiety symptoms in epilepsy patients are evaluated by the scales and inventories established. A quantitative measure of recurrent and severity of mood symptomatology experienced by them can be recorded by these tests [13, 14]. Neuropsychological functions in TLE patients can also be influenced by antagonist effects such as underlying injury, mood disorders, and AEDs treatment. In this regard, neuropsychological assessments are substantial to comprehend better the impact of those variables on the lives of patients. The interaction between laterality of temporal lobe epilepsy, usage of AEDs, depressive symptoms, and neuropsychological functions remains to be elucidated. For a better comprehension of this association, we proposed a systematic and comprehensive literature review searching for the outcomes of the correlation between these aspects in surgical patients.

## Methods

A systematic review was conducted using the methodology outlined in the Cochrane Handbook for Systematic Reviewers [15]. The data were reported following the Preferred Reporting Items for Systematic Reviews and Meta-Analyses [16]. The review protocol was registered under the number CRD42019104443 in the International Register of Prospective Systematic Reviews.

### Database search

A literature search was performed in PubMed/Medline, Web of Science, and PsycNET to obtain data from inception until January 2019. The following keywords and medical subject headings (MeSH) were used in the search strategy: (“epilepsy”, “temporal lobe” OR “epilepsy” AND “temporal” AND “lobe” OR “temporal lobe epilepsy” OR “temporal” AND “lobe” AND “epilepsy” AND “depressive disorder” OR “depressive” AND “disorder” OR “depressive disorder” OR “depression” OR “depression” OR “anxiety” OR “anxiety”. The strategies for other databases are available on request. Articles published in all languages were included. The bibliography of the included articles was screened manually. The titles and abstracts of all studies identified in the search based on the abovementioned terms and MeSH were evaluated independently by two authors (E.L.C. and F.S.S.). Disagreements were resolved by consensus or by a third reviewer (G.R.).

### Eligibility criteria

The inclusion criteria for the present review were as follows: articles without language restriction; diagnosis of epilepsy by ECG (Electrocardiogram) or MRI (Magnetic Resonance Imaging); Psychiatric diagnosis (by scales, interviews, diagnostic manuals – DSM and ICD -10); study design (case series with more than ten patients, retrospective and prospective, clinical, human); unilateral or bilateral temporal lobe epilepsy; surgical patients.

Studies of systematic reviews, letters, and experimental studies; children under 16 years; patients with dysphoria; patients with generalized epilepsy (multiple foci); non-surgical patients were the exclusion criteria. Figure 1 displayed a flowchart of study selection and inclusion.

**Figure 1.**
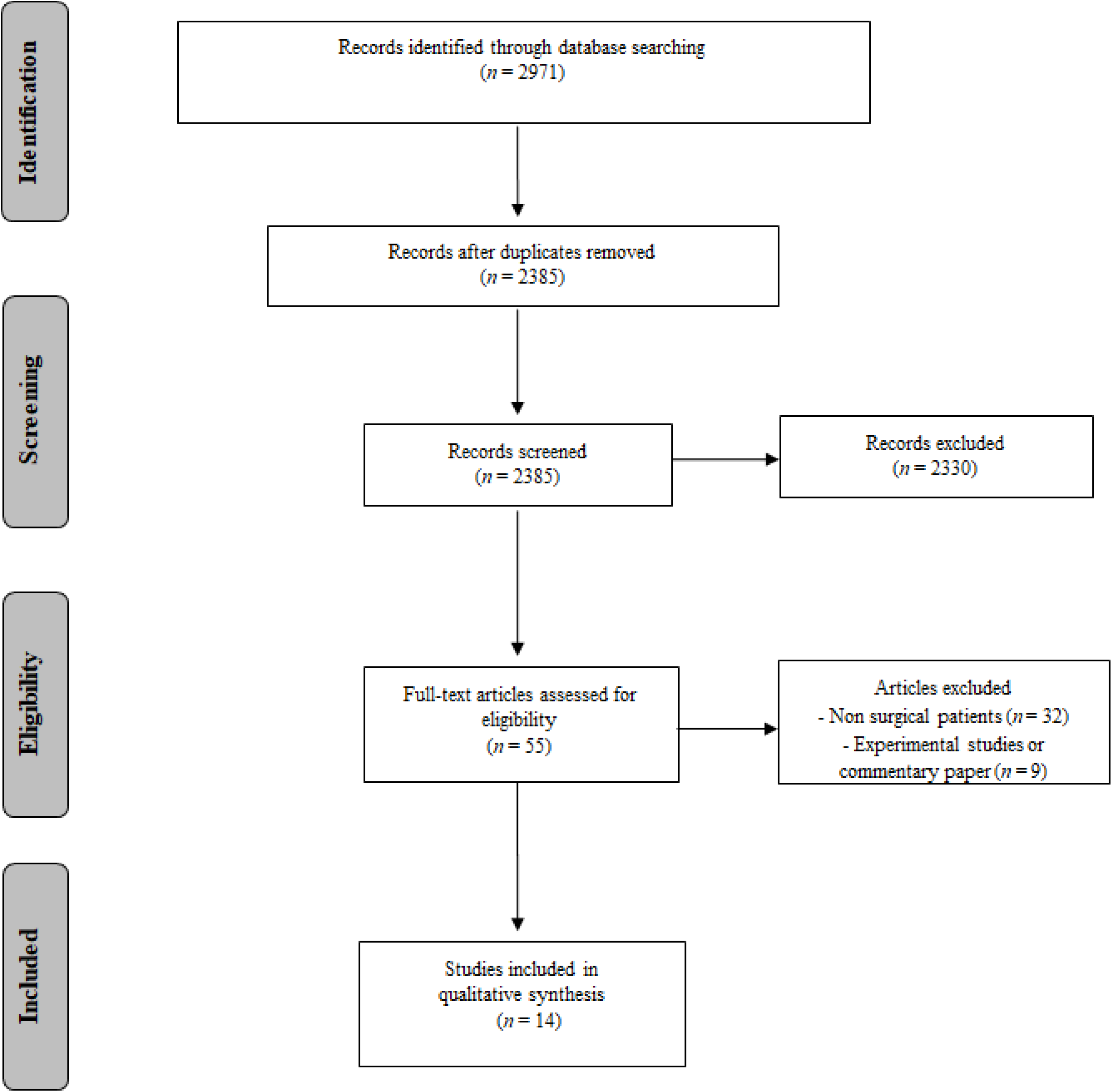
Summary of evidence search and study selection.

### Data extraction

The databases were investigated, and duplicate entries were eliminated. The full-text articles whose abstracts did not provide sufficient information regarding the inclusion and exclusion criteria were selected for evaluation. In the second phase, the same reviewers independently evaluated the full text of these articles and made their selection based on the eligibility criteria. Data on the following were collected: the number of patients, method of psychiatric diagnosis, symptoms diagnosis criteria, diagnostic scales, neuropsychological tests, report of the number of antiepileptic drugs, lateralization of the epileptic focus, individual depression symptoms, lateralized depression symptoms, individual anxiety symptoms, lateralized anxiety symptoms, and the design/classification of evidence.

## Results

The search retrieved 2971 potentially relevant citations from the electronic databases. By removing duplicate titles, 55 articles were eliminated. After screening titles and abstracts, only 14 articles met the inclusion criteria (Fig. 1). The clinical design approach included prospective (28.5%), retrospective (28.5%), cross-sectional follow-up (21.4%), and cohort (7.14%). Most of them were single-center studies (85.7%).

Overall, 14 surgical studies with TLE were evaluated and DSM/SCID was the main method used for psychiatric diagnosis (PD) (35.7%). However, about 50% of the studies did not employ any kind of PD. Moreover, the neuropsychological evaluation was not explored by most of the studies (71.5%). From 11 studies that portrayed clinical cases of lateralization of the epileptic focus, most of them were found to occur in the left hemisphere (483 patients) (Table 1). Nine studies were evaluated for individual depressive diagnosis and five to anxiety, therefore, from the data analyzed in both situations, no diagnosis was representative in preoperative and postoperative cases (p < 0.05) (Table 2).

**Table 1.** Clinical characteristics of surgical patients with Temporal Lobe Epilepsy related to the diagnostic and treatment.

| Study | N/(%) |
| --- | --- |
| **Sample (n = 14)** |  |
| Mean (SD) | 103,14 (±107,44) |
| **Design** |  |
| Cross-sectional follow-up | 3 (21.4) |
| Prospective | 6 (28.5) |
| Retrospective | 4 (28.5) |
| Cohort | 1 (7.14) |
| **Trail** |  |
| Single center | 12 (85.7) |
| Multi center | 2 (14.2) |
| **Method of Psychiatric Diagnosis** |  |
| DSM/SCID | 5 (35.7) |
| CIDI | 2 (14.3) |
| None | 7 (50) |
| **Neuropsychological Evaluation** |  |
| Yes | 4 (28.5) |
| No | 10 (71.5) |
| **Lateralization of epileptic focus (n = 11)** |  |
| Right | 457 |
| Left | 483 |
| Bilateral | 2 |
| **Use of Antiepileptic Drugs** |  |
| Reported | 6 (42.8) |
| Not Reported | 8 (57.2) |

*DSM, Diagnostic and Statistical Manual of Mental Disorders; SCID, Structured Clinical Interview for DSM; CIDI, Composite International Diagnostic Interview*

**Table 2.**
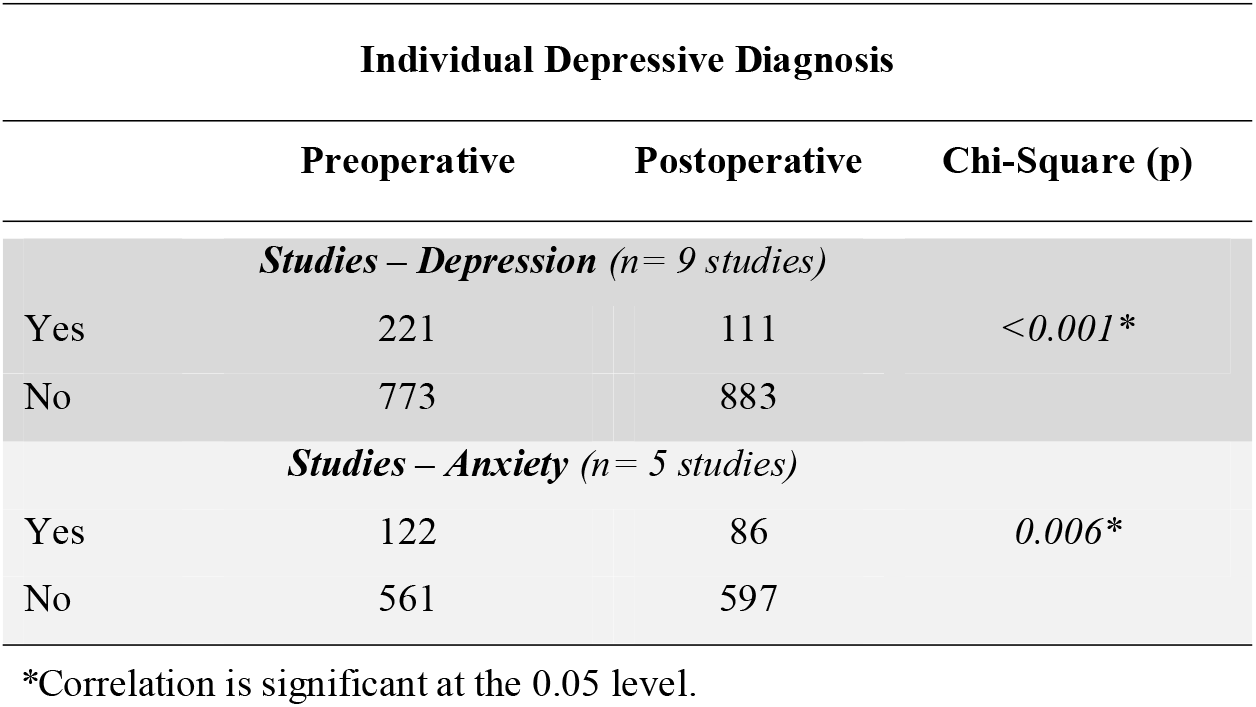
Clinical characteristics related to the lateralization of epileptic focus and depression and anxiety symptoms of surgical patients with Temporal Lobe Epilepsy.

| Individual Depressive Diagnosis |  |  |  |
| --- | --- | --- | --- |
|  | Preoperative | Postoperative | Chi-Square (p) |
|  | *Studies – Depression (n= 9 studies)* |  |  |
| Yes | 221 | 111 | <0.001* |
| No | 773 | 883 |  |
|  | *Studies – Anxiety (n= 5 studies)* |  |  |
| Yes | 122 | 86 | 0.006* |
| No | 561 | 597 |  |

\*Correlation is significant at the 0.05 level.

Regarding the types of treatment using antiepileptic drugs, most studies failed to report the treatment utilized (57.2%). More details about each study evaluated above are summarized in Tables 3 and 4.

**Table 3.**
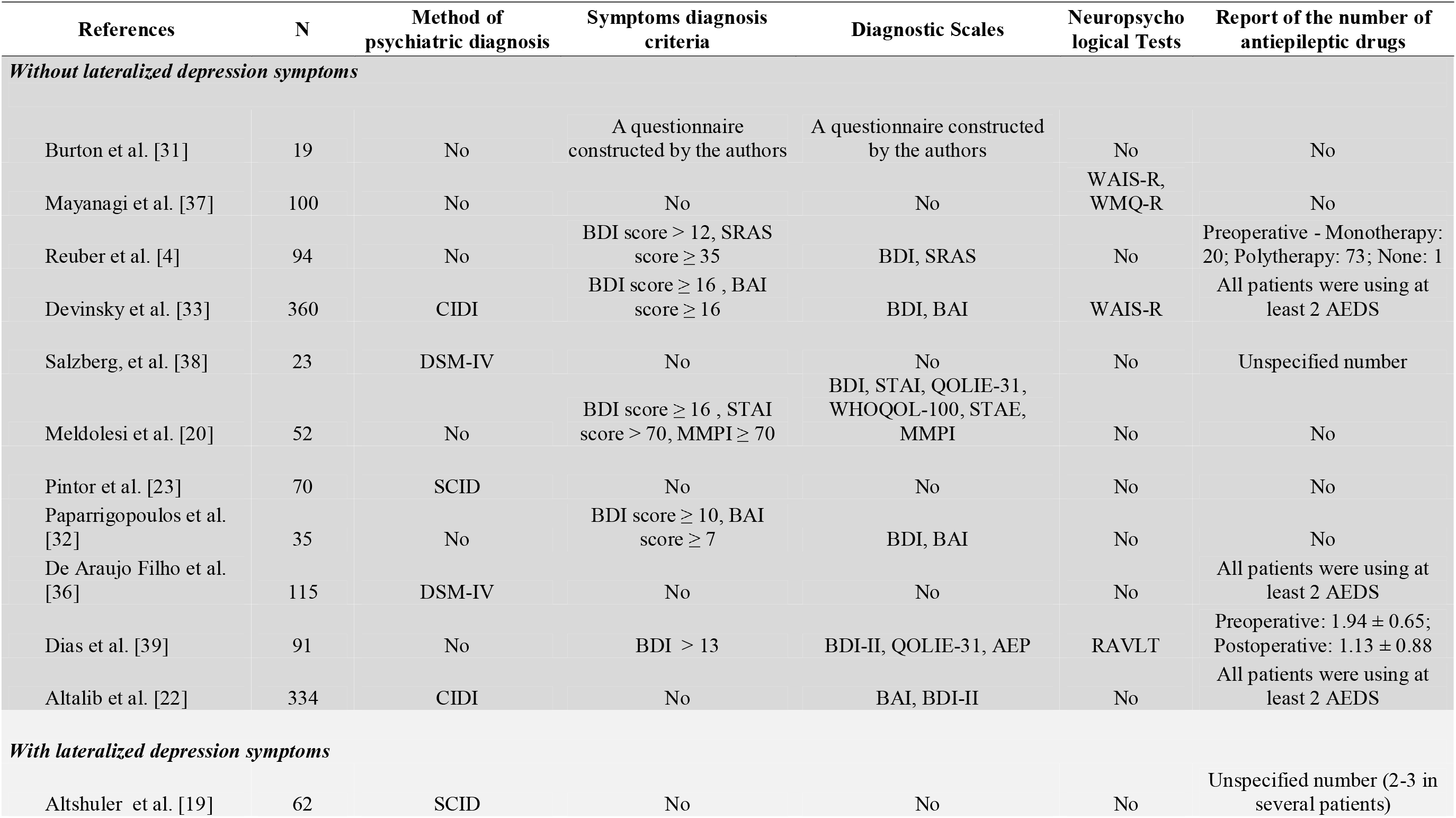
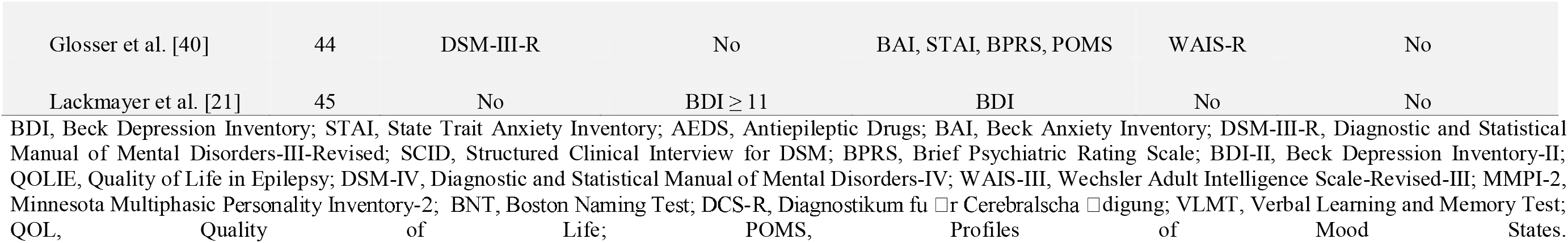
Clinical characteristics related to the diagnostic and treatment of surgical patients with Temporal Lobe Epilepsy.

**Table 4.**
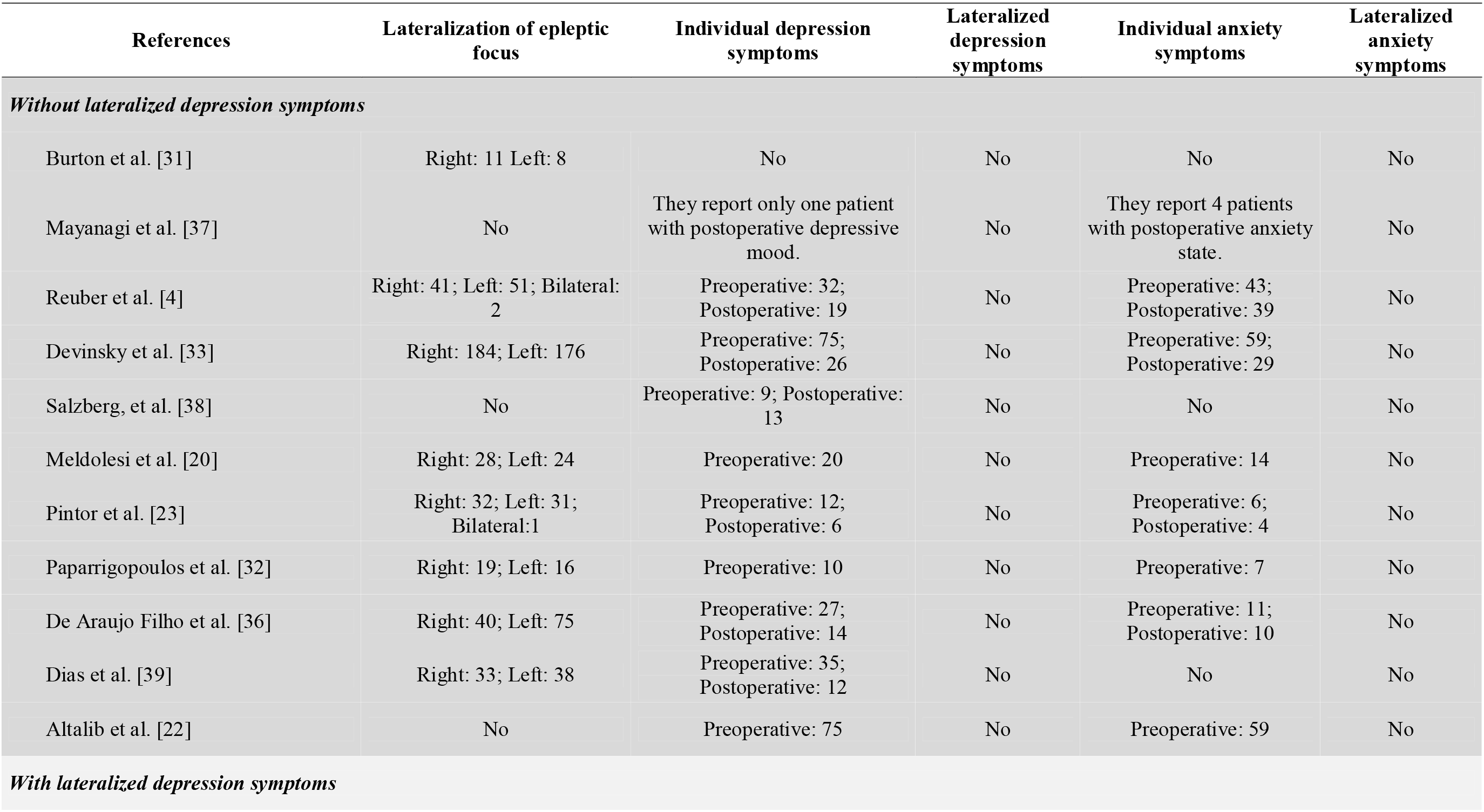
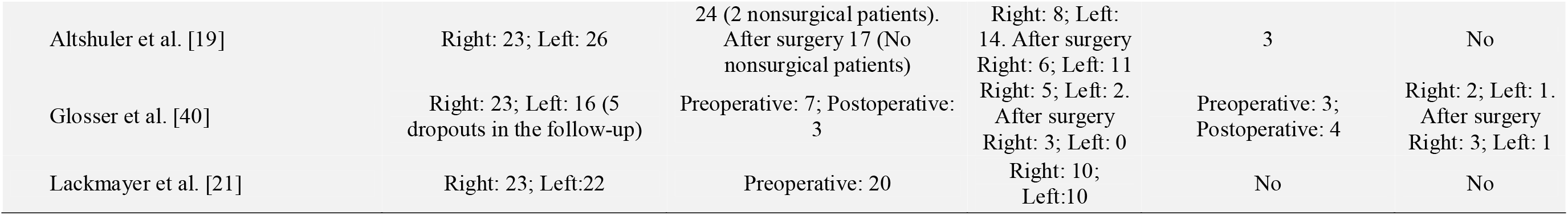
Clinical characteristics related to the lateralization of epileptic focus and depression and anxiety symptoms of surgical patients with Temporal Lobe Epilepsy.

## Discussion

Several beneficial outcomes, as reduced seizure frequency, improved quality of life, and mood dysfunction, have been known to be associated with surgical procedures for treating refractory epilepsy [17, 18]. Temporal lobectomy could be a resolution for depression and anxiety symptoms in epileptic patients since the limbic system play a crucial role in increasing the risk of these conditions [19]. Studies documented that for the patients with seizure focus in the temporal lobe, depression symptoms are more frequent as compared to those with an epileptic focus in extratemporal lobe regions, and the epilepsy surgery could resolve the depression symptoms in some of the patients [19]. Reduction in depression symptoms post-surgery has been reported by some studies, especially when patients presented decreased seizure frequency [4]. Contrary to this finding, Meldolesi and colleagues showed that though there was a reduction in anxiety symptoms after epilepsy surgery, depression persisted, even with the decline of seizure frequency in the majority of the patients [20]. Studies also indicated that the diagnosis of depression prior to the surgery did not affect the surgical outcome for seizure control [21,22]. However, Pintor et al. could relate the prevalence of psychiatric symptoms before surgery with the occurrence of psychiatric conditions post-surgery [23]. Furthermore, studies have highlighted the manifestation of anxiety in most of the epilepsy surgery candidates [24]. Anxiety can remain during the first year post-surgery [4] and can depend on the seizure control after surgery [25].

Though a number of treatments are available to reduce seizure frequency, its effect on depression or anxiety symptoms in epileptic patients should also be verified. The AEDs used for the treatment were not mentioned in most of the studies, considered for the present review. However, it is an important aspect since antiepileptic drugs may be responsible for an increase or decrease in depression and anxiety symptoms in epileptic patients [26]. Moreover, some antidepressant drugs may contribute to the alteration of seizure frequency [27]. Besides, it is also important to report any change in the number of AEDs prescribed to the patients after surgery. Prayson and colleagues demonstrated that the quantity of AEDs was reduced in only 28% of the patients after surgery, whereas 58% of patients continued with the same AEDs after surgery [28].

In the present review, the majority of the evaluated studies have shown lateralization of epileptic focus occurred in the left hemisphere. However, no clear consensus is present regarding the association of depression lateralization and epileptic focus. Some studies depicted that depression occurs more frequently after lesions or seizure focus in the left hemisphere [29, 30]. According to Burton and Labar [31], increased depression was reported by patients who were subjected to left temporal lobectomy in comparison to the patients with right temporal lobectomy [31]. Besides, a correlation between the severity of depression and the extension of hippocampus and amygdala resection, especially in left temporal lobectomy, was established by Paparrigopoulos et al. [32]. Furthermore, surgical laterality and location can be predisposing factors for psychological outcomes in epileptic patients post-surgery. Prayson et al. [28] elucidated more significant depression and anxiety symptoms among patients with left temporal lobe epilepsy before surgery as compared to the patients with left frontal lobe epilepsy. On the other hand, Devinsky et al. [33] demonstrated no association between the presence or absence of depression and anxiety and the lateralization or localization of the seizure onset before surgery. Besides, Manchanda et al. also failed to establish any correlation between the laterality of seizure focus and depression symptoms [24].

In this present systematic review, the evaluated data related to individual depressive diagnosis and anxiety, in both situations, no diagnosis was representative in preoperative and postoperative cases. Any kind of psychiatric diagnosis or neuropsychological evaluation was not executed by most of the evaluated studies in this review. The included studies that performed PD the primary method utilized was DSM/SCID. Different methods of depression diagnosis may lead to distinct conclusions and create hurdles in generalizing the outcomes [34]. Besides, some studies have recognized the preoperative depression diagnosis as potential complications in the postoperative seizure outcome of patients with refractory epilepsy [35]. De Araujo Filho and coauthors also demonstrated the absence of depression diagnosis pre or postoperative was associated with favorable seizure outcomes after surgery [36].

## Conclusions

The prevalence of psychiatric disorders among refractory epileptic patients undergoing surgical procedures has been documented by various studies. However, no clear association has been elucidated, so far, between lateralization of seizure focus and the existence of depression and anxiety conditions. Surgery is considered the best treatment option for refractory epileptic patients. Determination of the risk of depression and anxiety symptoms in epileptic patients before surgical procedures is important to estimate the efficacy of surgery in the psychiatry episodes and its relation to seizure control. Besides, identification of these symptoms prior to surgery is vital to distinguish those patients who will require a longer specific psychological surveillance after surgery. Thus, rigorous pre- and postoperative evaluation of psychiatry conditions is essential in patients with refractory epilepsy candidates for surgery.

## Data Availability

not applicable

## Declarations

### Ethics approval

not applicable.

### Consent to participate

not applicable.

### Consent for publication

not applicable.

### Availability of data and material

not applicable.

### Code availability

not applicable.

## ACKNOWLEDGEMENTS

JCC is funded by CNPq (research productivity scholarship). JCC is supported by Conselho Nacional de Desenvolvimento Científico e Tecnológico – (CNPq) Brazil grant PQ 307372/2015-4. Our study was supported by the following grants: CNPq (Conselho Nacional de Desenvolvimento Científico e Tecnológico); Coordenação de Aperfeiçoamento de Pessoal de Nível Superior (CAPES).

## Funding Sources

This research did not receive any other specific grant from funding agencies in the public, commercial, or not-for-profit sectors.

**Abbreviations**
TLE: Temporal Lobe Epilepsy
BDI: Beck Depression Inventory
STAI: State Trait Anxiety Inventory
AEDS: Antiepileptic Drugs
BAI: Beck Anxiety Inventory
DSM-III-R: Diagnostic and Statistical Manual of Mental Disorders-III-Revised
SCID: Structured Clinical Interview for DSM
BPRS: Brief Psychiatric Rating Scale
BDI-II: Beck Depression Inventory-II
QOLIE: Quality of Life in Epilepsy
DSM-IV: Diagnostic and Statistical Manual of Mental Disorders-IV
WAIS-III: Wechsler Adult Intelligence Scale-Revised-III
MMPI-2: Minnesota Multiphasic Personality Inventory-2
BNT: Boston Naming Test
DCS-R: Diagnostikum fu □r Cerebralscha □digung
VLMT: Verbal Learning and Memory Test
QOL: Quality of Life
POMS: Profiles of Mood States

## References

1. Kwan P, Brodie MJ (2000) Early identification of refractory epilepsy. N Engl J Med. 342:314–19.

2. Löscher W (2005) How to explain multidrug resistance in epilepsy? Epilepsy currents 5: 107–12.

3. Blair RD (2012) Temporal lobe epilepsy semiology. Epilepsy research and treatment 2012: 751510.

4. Reuber M, Andersen B, Elger C.E., Helmstaedter C (2004) Depression and anxiety before and after temporal lobe epilepsy surgery. Seizure 13: 129-135.

5. Butler C, Zeman AZ (2005) Neurological syndromes which can be mistaken for psychiatric conditions. J Neurol Neurosurg Psychiatry 76: i31–i38

6. Sadler RM (2006) The syndrome of mesial temporal lobe epilepsy with hippocampal sclerosis: clinical features and differential diagnosis. Advances in Neurology 97:27–37.

7. Brandt C, Schoendienst M, Trentowska M, May TW, Pohlmann-Eden B, Tuschen-Caffier B, et al. (2010) Prevalence of anxiety disorders in patients with refractory focal epilepsy--a prospective clinic based survey. Epilepsy Behav. 17:259–263.

8. Epps SA, Weinshenker D (2013) Rhythm and blues: animal models of epilepsy and depression comorbidity. Biochemical pharmacology 85: 135–46.

9. Manchanda R, Schaefer B, McLachlan RS, Blume WT (1992) Interictal psychiatric morbidity and focus of epilepsy in treatment refractory patients admitted to an epilepsy unit. The American Journal of Psychiatry 149:1096–98

10. Swinkels WAM, van Emde Boas W, Kuyk J, Van Dyck R, Spinhoven P (2006) Interictal depression, anxiety, personality traits, and psychological dissociation in patients with temporal lobe epilepsy (TLE) and extra-TLE. Epilepsia 47:2092–103.

11. Foong J, Flugel D (2007) Psychiatric outcome of surgery for temporal lobe epilepsy and presurgical considerations. Epilepsy Research 75:84–96.

12. Mansouri A, Fallah A, Valiante TA (2012) Determining surgical candidacy in temporal lobe epilepsy. Epilepsy research and treatment 706917.

13. Brooks J, Baker GA, Aldenkamp AP (2001) The A-B neuropsychological assessment schedule (ABNAS): the further refinement of a patientbased scale of patient-perceived cognitive functioning. Epilepsy Res. 43:227–37

14. Kwon OY, Park SP (2014) Depression and anxiety in people with epilepsy. Journal of clinical neurology 10: 175–188.

15. Higgins J, Thomas J (2011) Cochrane handbook for systematic reviews of interventions.

16. Moher D, Liberati A, Tetzlaff J, Altman DG; PRISMA Group (2009) Preferred reporting items for systematic reviews and meta-analyses: the PRISMA statement. PLoS Med. 6:e1000097.

17. Kellett MW, Smith DF, Baker GA, Chadwick DW (1997) Quality of life after epilepsy surgery. Journal of Neurology, Neurosurgery, and Psychiatry 63:52–58.

18. Alonso NB, Silva TI, Westphal-Guitti AC, Azevedo AM, Caboclo LOSF, Sakamoto AC, Yacubian EMT (2006) Quality of life related to surgical treatment in patients with temporal lobe epilepsy due to mesial temporal sclerosis. Journal of Epilepsy and Clinical Neurophysiology 12: 233-241.

19. Altshuler LL, Devinsky O, Post RM, Theodore W (1990) Depression, anxiety, and temporal lobe epilepsy: laterality of focus and symptoms. Archives of Neurology 47:284–288.

20. Meldolesi GN, Di Gennaro G, Quarato PP, Esposito V, Grammaldo LG, Morosini P, Cascavilla I, Picardi A (2007) Changes in depression, anxiety, anger, and personality after resective surgery for drug-resistant temporal lobe epilepsy: a 2-year follow-up study. Epilepsy Res. 77:22–30.

21. Lackmayer K, Lehner-Baumgartner E, Pirker S, Czech T, Baumgartner C (2013) Preoperative depressive symptoms are not predictors of postoperative seizure control in patients with mesial temporal lobe epilepsy and hippocampal sclerosis. Epilepsy Behav. 26: 81-86.

22. Altalib HH, Berg AT, Cong X, Vickrey BG, Sperling MR, Shinnar S, et al. (2018) Presurgical depression and anxiety are not associated with worse epilepsy surgery outcome five years postoperatively. Epilepsy Behav. 83:7–12.

23. Pintor L, Bailles E, Fernandez-Egea E, et al. (2007) Psychiatric disorders in temporal lobe epilepsy patients over the first year after surgical treatment. Seizure 16:218–25.

24. Manchanda R, Schaefer B, McLachlan RS, Blume WT, Wiebe S, Girvin JP, Parrent A, Derry PA (1996) Psychiatric disorders in candidates for surgery for epilepsy. Journal of Neurology, Neurosurgery, and Psychiatry 61:82–89.

25. Mattson P, Tibblin B, Kihlgreen M, Kumlien E (2005) A prospective study of anxiety with respect to seizure outcome after epilepsy surgery. Seizure 14:40–5.

26. Mula M, Schmitz B (2009) Depression in Epilepsy: Mechanisms and Therapeutic Approach. Ther Adv Neurol Disord. 2: 337–44.

27. Cardamone L, Salzberg MR, O’Brien TJ, Jones NC (2013) Antidepressant therapy in epilepsy: can treating the comorbidities affect the underlying disorder? Br J Pharmacol. 168:1531–54.

28. Prayson BE, Floden DP, Ferguson L, Kim KH, Jehi L, Busch RM (2017) Effects of surgical side and site on psychological symptoms following epilepsy surgery in adults. Epilepsy & Behavior 68:108–114.

29. Davidson R (1993) Parsing affective space: perspectives from neuropsychology and psychophysiology. Neuropsychology 7:464–475.

30. Mendez MF, Taylor JL, Doss RC, Salguero P. (1994) Depression in secondary epilepsy: relation to lesion laterality. Journal of Neurology, Neurosurgery & Psychiatry. 57: 232-33.

31. Burton LA, Labar D. Emotional status after right vs. left temporal lobectomy. Seizure. 1999;8:116–9.

32. Paparrigopoulos T, Ferentinos P, Brierley B, Shaw P, David AS (2008) Relationship between post-operative depression/anxiety and hippocampal/amygdala volumes in temporal lobectomy for epilepsy. Epilepsy Res. 81:30–5.

33. Devinsky O, Barr WB, Vickrey BG, Berg AT, Bazil CW, Pacia SV, et al. (2005) Changes in depression and anxiety after resective surgery for epilepsy. Neurology. 65:1744–9.

34. Fiest KM, Dykeman J, Patten SB, Wiebe S, Kaplan GG, Maxwell CJ, Bulloch AG, Jette N (2013) Depression in epilepsy: a systematic review and meta-analysis. Neurology. 80:590–9.

35. Kanner AM (2006) Depression and epilepsy: a new perspective on two closely related disorders. Epilepsy Curr. 6:141–6.

36. de Araújo Filho GM, Gomes FL, Mazetto L, Marinho MM, Tavares IM, Caboclo LO et al. (2012) Major depressive disorder as a predictor of a worse seizure outcome one year after surgery in patients with temporal lobe epilepsy and mesial temporal sclerosis. Seizure 21:619–23.

37. Mayanagi Y, Watanabe E, Nagahori Y, Nankai M (2001) Psychiatric and neuropsychological problems in epilepsy surgery: analysis of 100 cases that underwent surgery. Epilepsia42:19–23.

38. Salzberg M, Taher T, Davie M, Carne R, Hicks RJ, Cook M (2006) Depression in temporal lobe epilepsy surgery patients: an FDG-PET study. Epilepsia 47:2125–30.

39. Dias R, Baliarsing L, Barnwal NK, Mogal S, Gujjar P (2017) Role of pre-operative multimedia video information in allaying anxiety related to spinal anaesthesia: a randomised controlled trial. Indian Journal of Anaesthesia 60: 843–7.

40. Glosser G, Zwil AS, Glosser DS, O’Connor MJ, Sperling MR (2000) Psychiatric aspects of temporal lobe epilepsy before and after anterior temporal lobectomy. J Neurol Neurosurg Psychiatry. 68:53–58.

